# Knowledge and attitudes of commercial motorcyclists on personal protective equipment in the crisis-affected Limbe and Tiko Health Districts of Cameroon toward injury prevention: A formative research study for a health education intervention

**DOI:** 10.1101/2025.05.06.25327113

**Authors:** Chrisantus Eweh Ukah, Nicholas Tendongfor, Alan Hubbard, Elvis A. Tanue, Rasheedat Oke, Nahyeni Bassah, Sandra I. McCoy, Larissa Kumenyuy Yunika, Claudia Ngeha Ngu, Rebecca Hemono, S. Ariane Christie, Dickson S. Nsagha, Alain Chichom-Mefire, Catherine Juillard

## Abstract

**Background:** In Cameroon, commercial motorcycle riders are essential for urban transportation; however, they face considerable health risks from road traffic accidents and workplace hazards. Despite the critical role of personal protective equipment (PPE) in reducing injury risks, riders often possess limited knowledge and attitudes towards PPE. This study aimed to assess the knowledge and attitudes of motorcycle riders in the Limbe and Tiko Health Districts.

**Methods:** A community-based cross-sectional study was conducted with 499 commercial motorcycle riders aged 18 and older in these districts from the 15^th^ of May 2024 to the 17^th^ of August 2024. Participants were selected through consecutive sampling at motorcycle pick-up locations after obtaining ethical approval from the University of Buea, with severely ill individuals excluded from the study. Trained research assistants administered structured questionnaires to gather data on socio-demographics, riding habits, and riders’ knowledge and attitudes regarding PPE use. Data analysis was performed using descriptive statistics with SPSS version 25, and Bloom’s Criteria was applied to classify participants’ knowledge as good or poor.

**Results:** The average age of the 499 riders was 32.2±7.6 years, all of whom were male, with 48.5% aged between 21-30 years. Only 32.1% held a valid motorcycle license, and 37.1% were internally displaced due to the ongoing socio-political crisis in the two English-speaking regions of Cameroon. The findings showed that 30.7% of riders had good knowledge of PPE, 26.1% displayed positive attitudes, while only 13.2% practiced good PPE usage. This study highlights significant deficiencies in knowledge, attitudes, and practices related to PPE among motorcycle riders in Limbe and Tiko Health Districts, underscoring the necessity for targeted health education interventions to enhance their understanding and usage of PPE, ultimately improving safety and reducing injuries among riders.

**What is known in the Topic:**

1. Commercial motorcycle riders are essential for urban transportation but face significant health risks from road traffic accidents and workplace hazards.
2. Personal protective equipment (PPE) is critical in reducing injury risks among motorcycle riders.
3. There is a general lack of awareness and understanding regarding the importance of PPE among motorcycle riders.

**What this study adds:**

1. This study provides empirical data on the knowledge, attitudes, and practices regarding PPE usage among commercial motorcycle riders specifically in the Limbe and Tiko Health Districts of Cameroon.
2. It highlights the low percentage of riders with good knowledge (30.7%), positive attitudes (26.1%), and proper practices (13.2%) related to PPE usage, indicating significant deficiencies in these areas.
3. The findings emphasize the urgent need for targeted health education interventions to improve understanding and usage of PPE among motorcycle riders, aiming to enhance safety and reduce injuries in this population.

## Background

Road traffic injury is the eighth leading cause of death globally and expected to be the 7^th^ leading cause of death by 2030[1]. It accounts for about 1.3 million death per year with 92% of this deaths occurring in the low and middle income countries including Cameroon [2]. In many low- and middle-income countries (LMICs), commercial motorcycle transport has become a vital means of livelihood and urban mobility, particularly in areas with inadequate public transportation systems[3]. In low and middle-income countries (LMICs), a significant proportion of road fatalities involve motorcyclists. Specifically, they account for about 39% in low-income countries and 34% in middle-income countries[4]. In some regions, like South-East Asia, motorcycles are the highest contributors to road accidents, accounting for about 34% of deaths. Additionally, a recent study indicated a notable increase in the proportion of fatalities involving motorcyclists in LMICs over the past decade, with a 44% increase reported[4]. In Cameroon, motorcycles are a popular mode of transport in both rural and urban settings, offering flexible and affordable services[5]. However, the rapid expansion of commercial motorcycle operations has been accompanied by a rise in road traffic injuries (RTIs), with motorcycle riders and their passengers accounting for a substantial proportion of road trauma cases presenting to health facilities [6].

Personal protective equipment (PPE), including helmets, protective jackets, gloves, and boots, plays a critical role in reducing the severity of injuries sustained during motorcycle crashes[7–10]. Despite the proven effectiveness of PPE in preventing fatalities and serious injuries, compliance with PPE use among commercial motorcycle riders in many African settings remains suboptimal. This has been attributed to factors such as limited awareness, negative attitudes, poor enforcement of road safety regulations, and economic constraints[11,12]. Understanding the knowledge and attitudes of commercial motorcycle riders toward PPE is essential for designing targeted interventions that promote safety behaviors. While previous studies in Cameroon have examined the epidemiology of motorcycle-related injuries, few have focused on behavioral determinants such as knowledge and attitudes regarding PPE, particularly in high-risk districts like Limbe and Tiko. These urban centers in the South-West Region experience high volumes of motorcycle traffic due to the fact that, they are the two least affected health districts by the ongoing socio-political crisis in the two English speaking regions (Northwest and Southwest) of Cameroon[13].

The Limbe and Tiko Health Districts, located in the Southwest Region of Cameroon, have been significantly affected by the ongoing socio-political crisis in the English-speaking regions of the country[14]. The crisis has disrupted socioeconomic activities, displaced populations, and strained healthcare services. In such a context, commercial motorcycle riding has become a primary livelihood strategy for many, including internally displaced persons (IDPs) and unemployed youth. Many of the displaced riders in this area have little knowledge on safety measures on the highway. Personal protective equipment (PPE), such as helmets, gloves, and protective clothing, is essential for mitigating these risks[15].

This study seeks to address this gap by assessing the knowledge and attitudes of commercial motorcycle riders toward the use of PPE in the Limbe and Tiko Health Districts. Findings from this study will inform evidence-based strategies to improve safety practices among riders and contribute to the broader efforts in reducing motorcycle-related injuries in Cameroon.

## MATERIALS AND METHODS

### Study Area

The study was conducted over three months from the 15^th^ of May 2024 to the 17^th^ of August 2024 in the Limbe and Tiko Health Districts of Cameroon’s Southwest Region, which is currently affected by socio-political unrest. As the administrative centers of their respective districts, Limbe and Tiko are known for their economic activities, including agriculture, commerce, and tourism—particularly in Limbe, which boasts attractions like the Limbe Botanic Garden and Wildlife Centre. The road networks in these districts are crucial for transportation and commerce, with a notable presence of motorcycle riders. Given the significance of transportation and the risks associated with road traffic, researching road traffic injury prevention among motorcycle riders in Limbe and Tiko is vital for addressing public health and safety issues in these areas.

### Study Design

The study adopted a community-based cross-sectional design to assess the knowledge, and attitudes of commercial motorcycle riders regarding PPE in the crisis-affected Limbe and Tiko health districts.

### Study Population

The target population for this study comprised commercial motorcycle riders aged 18 and older who have been active in the Limbe and Tiko health districts for at least the past six months. These riders were specifically chosen due to their significant exposure to road traffic hazards and their potential to benefit from health education initiatives. In total, 499 commercial motorcycle riders participated in the study.

### Inclusion and Exclusion Criteria

The study included commercial motorcycle riders aged 18 years and above who had been operating in the Limbe or Tiko health districts for a minimum of six months and provided their consent to participate. Riders who faced language barriers or health limitations that prevented their participation were excluded from the study. This selection criteria ensured that the participants were well-acquainted with the local environment and could effectively engage with the study’s objectives.

### Sampling Method

A multistage sampling technique was employed to recruit participants for the study. The Limbe and Tiko health districts were divided into clusters based on geographical and administrative boundaries, as these areas are home to a significant population of commercial motorcycle riders. In the first stage, 80% of the motorcycle pick-up points were randomly selected from each district. Specifically, the Limbe health district, which has 98 motorcycle pick-up points, yielded 79 points (80% of 98), while the Tiko health district, with 34 pick-up points, contributed 27 points (80% of 34). In the second stage, major motorcycle parking areas or rider association meeting points within each selected cluster were identified as key sites for recruitment. At each of these points, riders were approached and recruited consecutively, ensuring a systematic and representative selection of participants for the study.

### Data Collection Tools and Procedures

Structured questionnaires were utilized to gather quantitative data regarding the knowledge, attitudes, and practices of motorcycle riders. To ensure the reliability and validity of the instruments, the questionnaires underwent a pre-testing phase in a similar setting prior to the main data collection. Data collection was carried out by eight trained research assistants who employed a mobile electronic data collection tool, specifically Kobo Collect. This approach facilitated efficient data entry and management. The research assistants administered the questionnaires by posing questions to the riders in English; where necessary, translations into Pidgin English were provided to ensure comprehension and accurate responses. The structured nature of the questionnaires allowed for systematic data collection, while the training of research assistants ensured consistency in the administration of the tool, thereby enhancing the overall quality of the data collected.

### Ethical Considerations

Ethical Clearance was obtained from the Institutional Review Board of the Faculty of Health Sciences of the University of Buea (2024/2490-03/UB/SG/IRB/FHS). Participation was voluntary, and written informed consent was sought from all respondents. Confidentiality and anonymity were maintained throughout the study. Administrative authorizations were obtained from the Department of Public Health of the University of Buea, South West Regional Delegation of Public Health and the District Health Services.

### Data Management and Analysis

The quantitative data collected through structured questionnaires were securely entered into a Kobo Collect database, which was accessible only to the principal investigator to ensure confidentiality and data integrity. The questions in the online questionnaire were made mandatory such that incompletely filled questionnaire could not be submitted. This eliminated the problem of missing data. The data analysis for this study on the knowledge and attitudes of commercial motorcycle riders toward personal protective equipment (PPE) was conducted using SPSS (Statistical Package for the Social Sciences). The analysis aimed to classify participants based on their overall knowledge and attitudes towards PPE, utilizing a structured scoring system. For the knowledge assessment, each participant’s responses to knowledge questions were scored dichotomously: a correct answer received a score of 1, while an incorrect answer received a score of 0. The total possible score for knowledge questions was calculated, and an 80% cutoff of the maximum obtainable score was established to classify participants into two categories[16]: those with overall good knowledge and those with poor knowledge. Participants achieving a score equal to or greater than the 80% threshold were categorized as having good knowledge, while those scoring below this threshold were classified as having poor knowledge. In assessing attitudes towards PPE, responses were scored on a five-point Likert scale, where “strongly disagree” was assigned a score of 0, “disagree” received a score of 1, “neutral” was scored as 2, “agree” as 3, and “strongly agree” as 4. The total attitude score for each participant was calculated by summing the scores across all attitude-related questions. Similar to the knowledge assessment, an 80% cutoff of the maximum obtainable score for attitudes was applied to classify participants into having overall positive attitudes or negative attitudes towards PPE.

Descriptive statistics were utilized to summarize the socio-demographic characteristics of the participants, as well as their knowledge and attitudes scores. Frequencies and percentages were calculated for categorical variables, while means and standard deviations were computed for continuous variables.

## RESULTS

### Socio-Demographic Variables of Commercial Motorcycle Riders

Regarding the socio-demographic characteristics of motorcycle riders **(Table 1),** 242 (48.5%) were within the age range 21-30 years and all the riders were males and 261 (52.3%) were single. For the highest level of education 291 (58.3%) had attended secondary school. A total of 270 (54.1%) riders’ monthly income was between 50,000–100,000 XAF and about half of them perceived their income as sufficient, 259 (51.9%). With respect to riding duration, the majority had been riding for 1–5 years, 308 (61.7%) and 297 (59.5%) reported primarily riding in both urban and rural areas. The proportion of riders who reported not having a valid motorcycle license was 339 (67.9%). Regarding smoking habits, 380 (76.2%), reported they do not smoke, while for alcohol consumption, the majority of riders reported drinking alcohol, 341 (68.3%). A total of 314 (62.9%) were not internally displaced persons. Finally, with respect to having a resting day, the majority of riders indicated they do have a resting day, 425 (85.2%).

**Table 1:** Socio-demographic variables of motorcycle riders.

| Variable | Category | Frequency | Percentage |
| --- | --- | --- | --- |
| Age group (years) | 21-30 | 242 | 48.5 |
|  | 31-40 | 192 | 38.5 |
|  | 41-50 | 57 | 11.4 |
|  | 50+ | 8 | 1.6 |
|  | **Total** | **499** | **100** |
|  | Male | 499 | 100 |
|  | **Total** | **499** | **100** |
| Marital status | Single | 261 | 52.3 |
|  | Married | 228 | 45.7 |
|  | Divorced | 10 | 2.0 |
|  | **Total** | **499** | **100** |
| Highest level of education | No formal | 19 | 3.8 |
|  | Primary | 149 | 29.9 |
|  | Secondary | 291 | 58.3 |
|  | Tertiary | 40 | 8.0 |
|  | **Total** | **499** | **100** |
| Average monthly income<br>(XAF1000) | <50 | 20 | 4.0 |
|  | 50-100 | 270 | 54.1 |
|  | 101-150 | 158 | 31.7 |
|  | 150+ | 51 | 10.2 |
|  | **Total** | **499** | **100** |
| Income perception | Insufficient | 220 | 44.1 |
|  | Sufficient | 259 | 51.9 |
|  | Largely sufficient | 20 | 4.0 |
|  | **Total** | **499** | **100** |
| Riding duration (years) | 1-5 | 308 | 61.7 |
|  | 11-15 | 45 | 9.0 |
|  | 15+ | 20 | 4.0 |
|  | 6-10 | 126 | 25.3 |
|  | **Total** | **499** | **100** |
| Riding location | Rural | 14 | 2.8 |
|  | Urban | 188 | 37.7 |
|  | Both | 297 | 59.5 |
|  | **Total** | **499** | **100** |
| Primary riding location | No | 102 | 20.4 |
|  | Yes | 397 | 79.6 |
|  | **Total** | **499** | **100** |
| Own a valid license | No | 339 | 67.9 |
|  | Yes | 160 | 32.1 |
|  | **Total** | **499** | **100** |
| Smoker | No | 380 | 76.2 |
|  | Yes | 119 | 23.8 |
|  | **Total** | **499** | **100** |
| Drink alcohol | No | 158 | 31.7 |
|  | Yes | 341 | 68.3 |
|  | **Total** | **499** | **100** |
| Internally displaced person | No | 314 | 62.9 |
|  | Yes | 185 | 37.1 |
|  | **Total** | **499** | **100** |
| Have a resting day | No | 74 | 14.8 |
|  | Yes | 425 | 85.2 |
|  | Total | 499 | 100 |

### Knowledge of Commercial Motorcycle Riders on Personal Protective Equipment

Regarding knowledge of commercial motorcycle rider on personal protective equipment (Table 3), a total of 380 (76.2%) knew that, the primary purpose of wearing a helmet was to protect the head in case of a road traffic crash and 204 (40.9%) knew that, the primary benefit of wearing gloves was to reduce the risk of hand injury in case of a road traffic crash of fall. Majority of the riders 369 (73.9%) knew that, the recommended shoes for commercial motorcycle riders was closed-toe-shoes and 388 (77.8%) knew that, the primary purpose of wearing eye glasses while riding was to protect the eyes from debris, dust or insects. A majority of the riders 320 (64.1%) correctly reported that, the correct recommendation of wearing was to wear it on every ride and 248 (49.7%) reported that their personal protective equipment should be inspected only when they notice a problem. A total of 219 (43.9%) reported that, the overall primary benefit of wearing personal protective equipment was to reduce the risk of injury in case of road traffic crash or fall.

**Table 3:** Knowledge of commercial motorcycle riders on personal protective equipment.

| Variable | Category | Frequency | Percentage |
| --- | --- | --- | --- |
| Primary purpose of wearing helmet | To attract passengers | 55 | 11.0 |
|  | To beautify the bike | 25 | 5.0 |
|  | To increase visibility | 39 | 7.8 |
|  | To protect the head in case of a road traffic crash or fall | 380 | 76.2 |
|  | **Total** | **499** | **100** |
| Primary benefit of gloves | I do not know | 20 | 4.0 |
|  | Improve grip on the steering | 89 | 17.8 |
|  | Make hands warm in cold weather | 186 | 37.3 |
|  | Reduce risk of hand injury in case of a road traffic crash or fall | 204 | 40.9 |
|  | **Total** | **499** | **100** |
| Recommended shoes for bike riders | Any shoe type is okay | 95 | 19.0 |
|  | Closed toe shoes | 369 | 73.9 |
|  | Sandals | 33 | 6.6 |
|  | Slippers | 2 | 0.4 |
|  | **Total** | **499** | **100** |
| Purpose of wearing eye wears | To attract passengers | 43 | 8.6 |
|  | To make the rider look profession | 36 | 7.2 |
|  | To protect the eyes from debris/dust and insects | 388 | 77.8 |
|  | To reduce visibility | 32 | 6.4 |
|  | **Total** | **499** | **100** |
| Recommendation for wearing PPE | Wear PPE in cold weathers to keep you warm | 25 | 5.0 |
|  | Wear PPE on every ride, regardless of distance or location | 320 | 64.1 |
|  | Wear PPE only during long rides | 92 | 18.4 |
|  | Wear PPE only in urban areas | 62 | 12.4 |
|  | **Total** | **499** | **100** |
| When to inspect PPE | Before every ride | 147 | 29.5 |
|  | Never | 12 | 2.4 |
|  | Once a year | 92 | 18.4 |
|  | When they notice a problem | 248 | 49.7 |
|  | **Total** | **499** | **100** |
| Primary benefit of wearing PPE | Attract passengers | 102 | 20.4 |
|  | Improve comfort while riding | 89 | 17.8 |
|  | Look responsible and professional | 89 | 17.8 |
|  | Reduce risk of injury in case of road traffic crash | 219 | 43.9 |
|  | **Total** | **499** | **100** |

### Motorcycle Riders’ Knowledge on the Different Types of PPE

Finally, regarding knowledge of riders on the different types of personal protective equipment, 422 (84.6%) correctly reported a helmet as a type of personal protective equipment (Figure 1).

**Figure 1:**
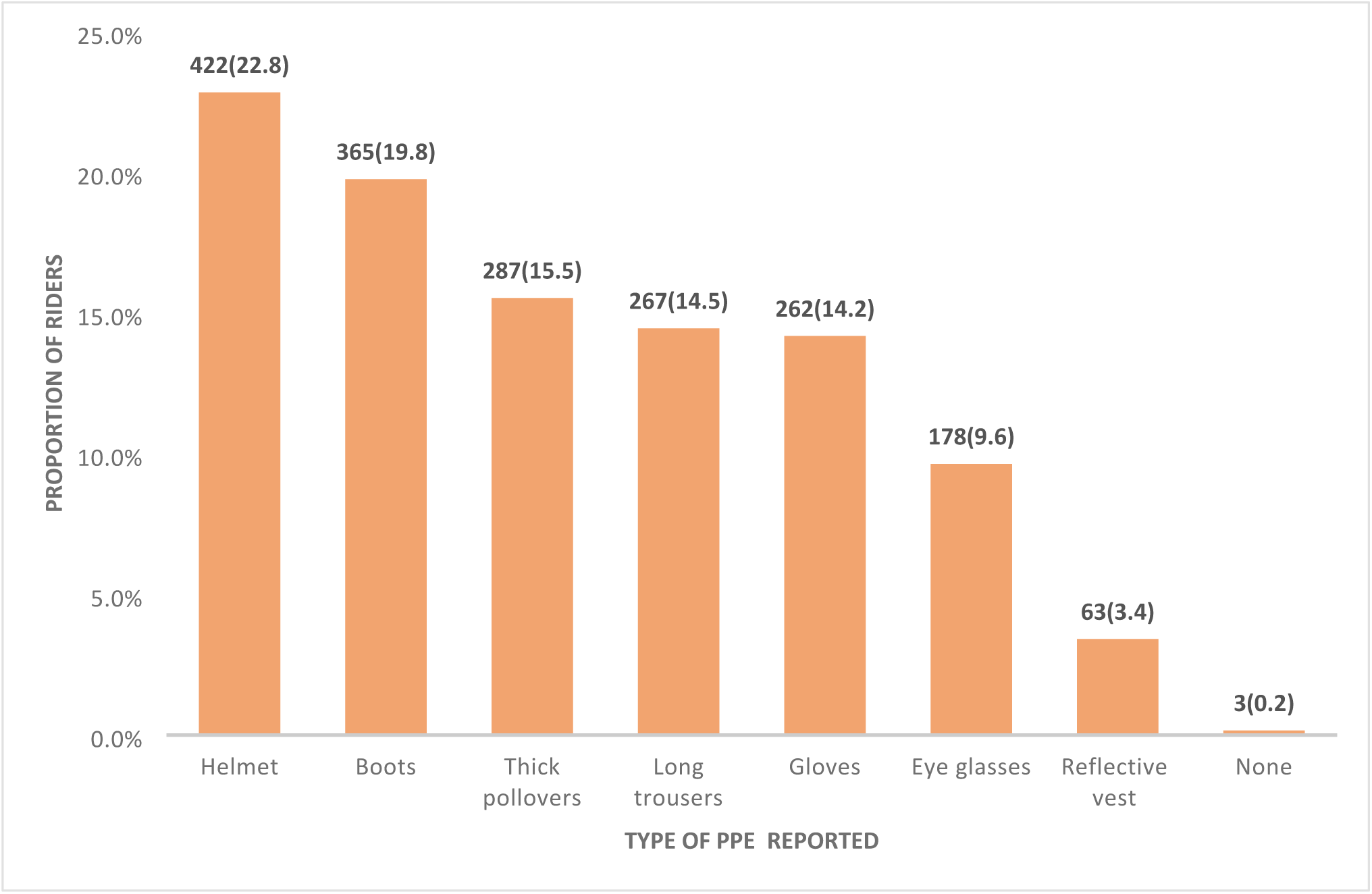
Motorcycle riders’ knowledge on the different types of personal protective equipment.

### Overall, Knowledge of Bike Riders on Personal Protective Equipment

Regarding the overall knowledge of riders on personal protective equipment (Figure 2), the composite scoring system was used. The Bloom’s Criteria was used to determine the cutoff point for good knowledge. A total of nine questions were asked with a maximum obtainable score of 13. The cutoff score following the Bloom’s Criteria at 80% of the maximum obtainable score was 10. Therefore, motorcycle riders who scored from 10 and above were classified as having good knowledge and those who scored below 10, were classified as having poor knowledge. Following this classification, the overall good knowledge of the riders on personal protective equipment in the two districts was 153 (30.7%). The Tiko Health District reported an overall good knowledge of 86 (43.2%) and Limbe Health District reported a good knowledge of 67 (22.3%).

**Figure 2:**
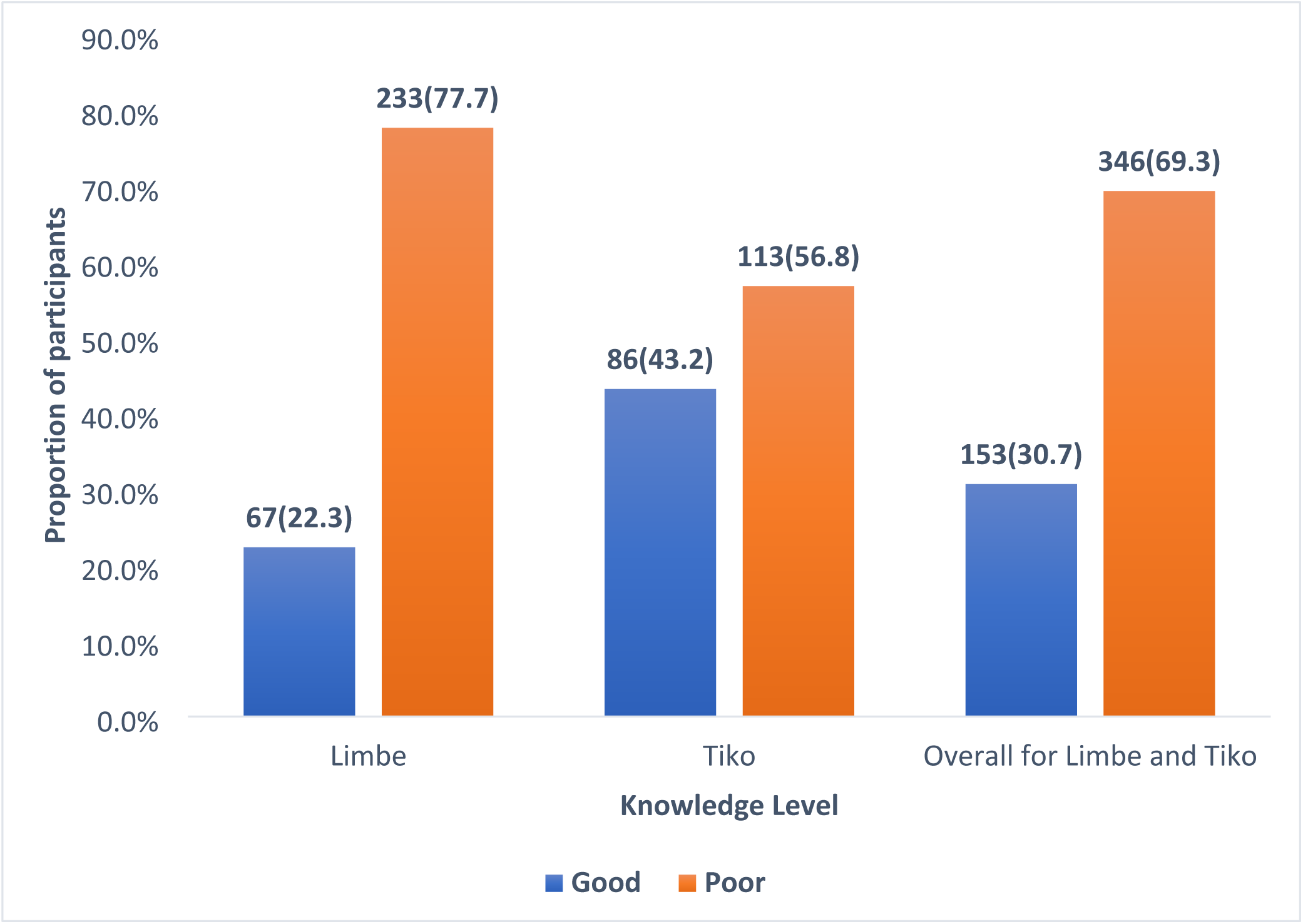
Overall knowledge of bike riders on personal protective equipment.

### Attitudes of Commercial Motorcycle Riders toward Personal Protective Equipment

With regards to the attitudes of commercial motorcycle riders toward personal protective equipment (Table 4), 216 (43.3) agreed that helmet was the most important personal protective equipment for motorcycle riders and 266 (53.3%) agreed that closed-toe-shoes were important for motorcycle riders. A total of 273 (54.7%) agreed that protective eye wears were important for motorcycle riders and 257 (51.5%) agreed that, they would wear personal protective equipment even if non wearing is not punishable by law. More than half of the riders 270 (54.7%) agreed feeling comfortable with the wearing of personal protective equipment while riding and 295 (59.1) agreed that wearing of personal protective equipment by commercial motorcycle riders is mandatory and not a personal choice. A total of 285 (57.1%) reported feeling comfortable with the enforcement of wearing personal protective equipment by riders and 304 (19.6%) agreed that, wearing personal protective equipment while riding was overall generally important.

**Table 4:** Attitudes of commercial motorcycle riders toward personal protective equipment.

| Variable | Strongly disagree | Disagree | Neutral | Agree | Strongly agree |
| --- | --- | --- | --- | --- | --- |
| Helmet is important to riders | 5(1.0) | 61(12.2) | 116(23.2) | 216(43.3) | 101(20.2) |
| Closed-toe-shoes are important to riders | 0(0) | 37(7.4) | 66(13.2) | 266(53.3) | 130(26.1) |
| Protective eye wears are important to riders | 6(1.2) | 44(8.8) | 86(17.2) | 273(54.7) | 90(18.0) |
| Protective eye wears are important to riders | 6(1.2) | 44(8.8) | 86(17.2) | 273(54.7) | 90(18.0) |
| will wear PPE even if not punishable | 7(1.4) | 60(12.0) | 110(22.0) | 257(51.5) | 65(13.0) |
| I feel comfortable when wearing PPE | 2(0.4) | 48(9.6) | 89(17.8) | 270(54.1) | 90(18.0) |
| wearing PPE is a necessity not a choice | 5(0.8) | 45(9.0) | 69(13.8) | 295(59.1) | 85(17.0) |
| feel comfortable with enforcement of PPE | 4(0.8) | 50(10.0) | 93(18.6) | 285(57.1) | 67(13.4) |
| I think the use of PPE is generally important | 2(0.4) | 27(5.4) | 68(13.6) | 304(60.9) | 98(19.6) |

### Overall Attitudes toward Personal Protective Equipment

Regarding the overall attitudes of commercial motorcycle riders toward personal protective equipment (Figure 3), the Bloom’s Criteria was used to analyze attitudes with a cutoff point set at 80% of the maximum obtainable score. A total of nine questions were asked in this section with a maximum obtainable score of 36. The options to the questions ranged from Strongly agree (a score of 4) to strongly disagree (a score of 0). A cut of score (80% of the maximum obtainable score) was 29. Therefore, commercial motorcycle riders who scored from 29 and above were classified as having positive attitudes and those who scored below 29 were classified as having negative attitudes. Overall, 130(26.1%) commercial riders had a positive attitude in both health districts. In the Tiko Health District, 93 (46.7%) had a positive attitude and 37 (12.3%) in the Limbe Health District.

**Figure 3:**
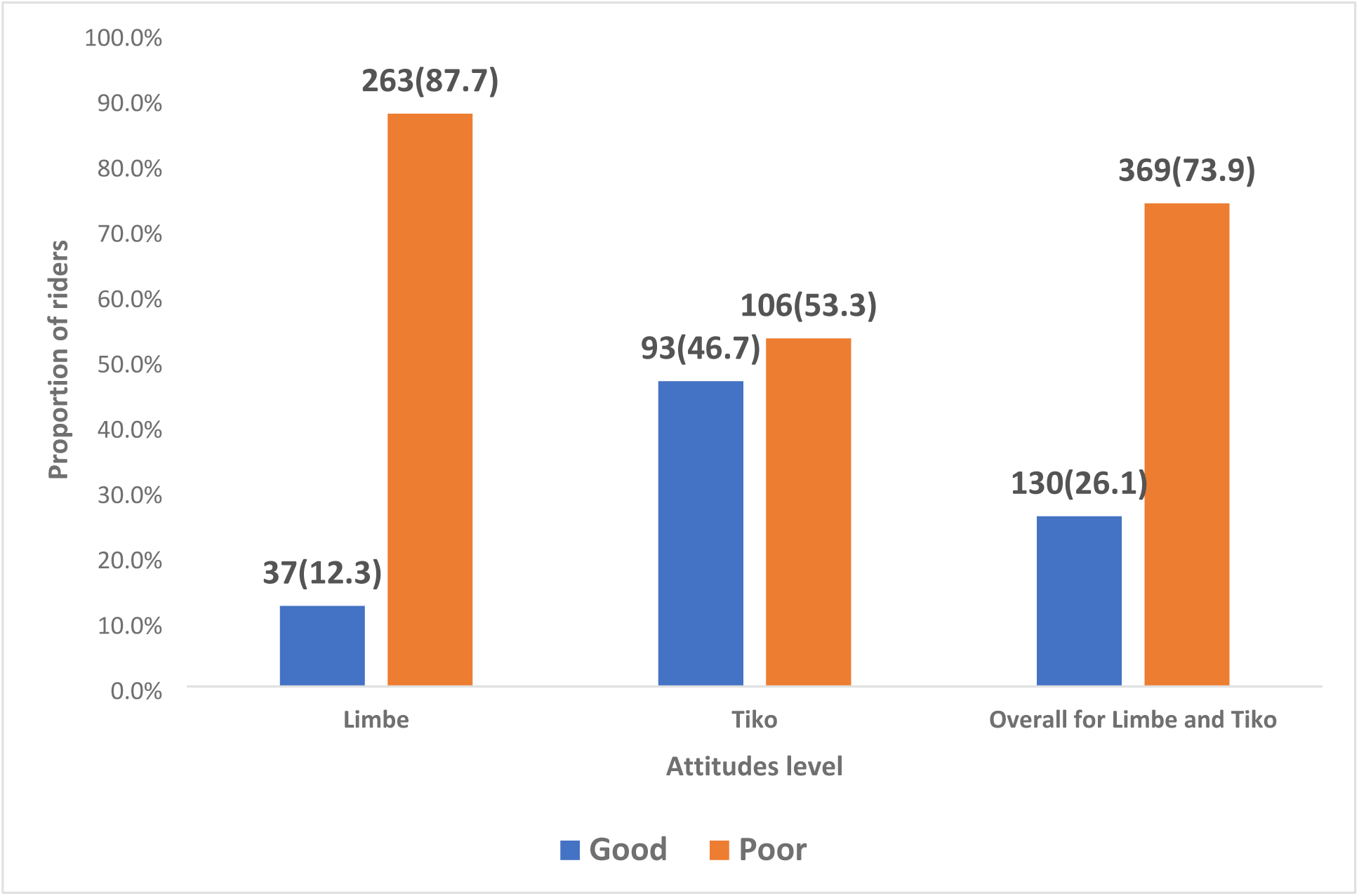
Overall attitudes of motorcycle riders toward personal protective equipment.

## DISCUSSION

This study aimed to assess the knowledge and attitudes on commercial motorcycle riders toward personal protective equipment.

### Knowledge of commercial motorcycle riders on Personal Protective Equipment

The results of this research indicate that the correct knowledge of commercial motorcycle riders regarding personal protective equipment (PPE) in the Limbe and Tiko Health Districts is relatively low, with an average of 30.7%. This low level of knowledge among motorcycle riders in these health districts is concerning as it indicates that, just about 3 in 10 riders have good knowledge on personal protective equipment. A closer examination reveals that the Tiko Health District exhibited a higher level of good knowledge at 43.2%, compared to only 22.3% in the Limbe Health District. These findings suggest a significant disparity in knowledge levels between the two districts. The lower level of knowledge exhibited by motorcycle riders in the Limbe Health District could be due to the fact that, 68.4% of riders with no formal education in this study were from the Limbe Health District. This higher proportion of uneducated riders might have contributed to the poorer knowledge in the Limbe Health District. Education has been shown to improve knowledge of motorcyclist on safety regulation including personal protective equipment[17]. In addition, 69.7% of internally displaced persons in this study were from the Limbe Health District. Internally displaced persons may engage in bike riding as a means of survival in their new environment without adequate training. This could also contribute to the lower knowledge level in the Limbe Health District.

For the implication of the findings, the low overall knowledge of PPE among motorcycle riders is concerning, as it suggests that a substantial number of riders may be at increased risk for injuries and fatalities associated with motorcycle accidents. According to the previous studies, motorcycle riders are particularly vulnerable to road traffic injuries[18–20], and the use of appropriate PPE can significantly reduce the severity of injuries sustained in accidents[15,21–23] The findings underscore the urgent need for targeted health education interventions that focus on increasing awareness and understanding of the importance of PPE among commercial motorcycle riders, especially in areas like Limbe where knowledge levels are critically low. Improved knowledge will enhance good practices[24].

This low good knowledge is similar to the 27% good knowledge reported among commercial motorcycle riders by Nasiru and his colleagues in Sokoto Metropolis, Northwestern Nigeria[25]. This is however different from what Adejumo Mumuni and colleagues in Oluyole Local Government Area, Ibadan, Nigeria reported that motorcycle riders had a good understanding of road safety regulation including personal protective equipment use[26]. This higher knowledge level reported in this place could be partly due to differences in the data analysis method. The study in Nigeria used the mean knowledge score as cut off point for good knowledge while in this study, the Bloom’s Criteria with cutoff point of 80% of the maximum obtainable score was used.

The disparities observed in this study could also be attributed to several factors. First, cultural differences and varying levels of enforcement of road safety regulations can influence knowledge and attitudes towards PPE[25]. In regions where helmet laws are strictly enforced, riders may be more aware of helmet use[27] but less informed about additional protective measures. Second, socioeconomic factors may also play a role; riders in areas with limited access to information or educational resources may exhibit lower knowledge levels.

This research highlights a critical gap in knowledge regarding personal protective equipment among commercial motorcycle riders in the Limbe and Tiko Health Districts. The findings indicate a pressing need for targeted health education interventions designed to enhance awareness and understanding of PPE. By addressing these gaps and building on the insights gained from this study, public health authorities can work towards reducing the incidence of motorcycle-related injuries and fatalities in these communities. This gab is being covered by the health education intervention we are implementing as part of this study to improve the knowledge of commercial motorcycle riders on personal protective equipment.

### Attitudes of commercial motorcycle riders towards personal protective equipment

The overall positive attitude toward PPE use across both districts was reported at 26.1%. In the Tiko Health District, a higher percentage of riders (46.7%) showed positive attitudes toward using PPE, whereas only 12.3% of riders in the Limbe Health District reported positive attitudes toward PPE adoption.

The disparity in attitudes between the two health districts can be attributed to several potential factors:

The low rate of positive attitudes in Limbe (12.3%) suggests that a significant number of commercial motorcycle riders may not be adopting basic safety measures, which could increase the risk of injuries or fatalities[28]. Motorcycle riders are particularly vulnerable to road traffic accidents[29], and the use of personal protective equipment (PPE), including helmets and reflective vests, is essential in mitigating these risks[30]. The findings suggest a need for enhanced safety measures to protect riders from avoidable injuries.

The need for targeted public health campaigns: Education campaigns could involve collaboration with local authorities, motorcycle rider associations, and community leaders to encourage the use of PPE and highlight its role in reducing injury rates. Finally, policy recommendation is another implication of the findings. To address the low adoption rates of PPE, policymakers should consider enforcing regulations that require commercial motorcycle riders to wear protective gear[31]. The significance of these results lies in their potential to inform future interventions and improve road safety for commercial motorcycle riders:

Enhancing Road Safety. The study underscores the need for targeted measures to increase PPE adoption among commercial motorcycle riders in both districts.

Previous studies on commercial motorcycle riders in other African countries have reported varying attitudes toward PPE. For example, a study in Nigeria found that only 28% of commercial motorcyclists used helmets, despite laws mandating their use[32]. Potential

The study’s results highlight significant disparities in the attitudes of commercial motorcycle riders toward personal protective equipment between the Tiko and Limbe Health Districts. While the results demonstrate a positive trend in Tiko, the overall low adoption rate in Limbe underscores the need for targeted public health interventions.

## Conclusion

This study highlights significant gaps in the knowledge, attitudes, and practices of commercial motorcycle riders in the crisis-affected Limbe and Tiko health districts of Cameroon regarding personal protective equipment (PPE) and injury prevention. The findings reveal that many riders lack adequate awareness and understanding of the importance of PPE, which exposes them to increased risk of injury. The insufficient levels of awareness and adoption of PPE among motorcycle riders emphasize the urgent need for targeted health education interventions to enhance rider safety. Without proper knowledge and positive attitudes towards safety practices, the likelihood of consistent and effective use of protective gear remains low. Engaging with local communities, stakeholders, and health authorities will be essential to develop culturally relevant strategies that resonate with riders’ experiences and realities.

## Data Availability

The data that support the findings of this study are available from the corresponding author upon reasonable request. Due to privacy and ethical considerations, the dataset contains sensitive information that cannot be made publicly available. However, summary statistics and findings are included in the manuscript. For inquiries regarding access to the data, please contact the corresponding author at

## Authors’ Contribution

**Conceptualization**: Chrisantus Eweh Ukah, Nicholas Tendongfor, Alan Hubbard, Elvis A. Tanue, Rasheedat Oke, Nahyeni Bassah, Sandra I. McCoy, Larissa Kumenyuy Yunika, Claudia Ngeha Ngu, S. Ariane Christie, Dickson S. Nsagha, Rebecca Hemono, Alain Chichom-Mefire, Catherine Juillard

**Data Curation**: Chrisantus Eweh Ukah, Nicholas Tendongfor, Alan Hubbard, Elvis A. Tanue, Rasheedat Oke, Nahyeni Bassah, Sandra I. McCoy, Larissa Kumenyuy Yunika, Claudia Ngeha Ngu, S. Ariane Christie, Dickson S. Nsagha, Rebecca Hemono, Alain Chichom-Mefire, Catherine Juillard

**Formal Analysis:** Chrisantus Eweh Ukah, Nicholas Tendongfor, Alan Hubbard, Elvis A. Tanue, Larissa Kumenyuy Yunika

**Funding Acquisition:** Ariane Christie, Sandra I. McCoy, Alain Chichom-Mefire, Catherine Juillard

**Investigation**: Chrisantus Eweh Ukah, Nicholas Tendongfor, Alan Hubbard, Elvis A. Tanue, Rasheedat Oke, Nahyeni Bassah, Sandra I. McCoy, Larissa Kumenyuy Yunika, Claudia Ngeha Ngu S. Ariane Christie, Dickson S. Nsagha, Alain Chichom-Mefire, Catherine Juillard

**Methodology:** Chrisantus Eweh Ukah, Nicholas Tendongfor, Alan Hubbard, Elvis A. Tanue, Rasheedat Oke, Nahyeni Bassah, Sandra I. McCoy, Larissa Kumenyuy Yunika, Claudia Ngeha Ngu, S. Ariane Christie, Dickson S. Nsagha, Alain Chichom-Mefire, Catherine Juillard

**Supervision:** Nicholas Tendongfor, Alan Hubbard, Catherine Juillard

**Validation:** Nicholas Tendongfor, Alan Hubbard, Elvis A. Tanue, Rasheedat Oke, Nahyeni Bassah, Ariane Christie,Catherine Juillard

**Visualization**: Chrisantus Eweh Ukah, Nicholas Tendongfor, Alan Hubbard, Elvis A. Tanue, Larissa Kumenyuy Yunika

**Writing – original draft:** Chrisantus Eweh Ukah, Nicholas Tendongfor, Alan Hubbard, Elvis A. Tanue, Rasheedat Oke, Nahyeni Bassah, Larissa Kumenyuy Yunika, Catherine Juillard

**Writing – Review and Editing:** Chrisantus Eweh Ukah, Nicholas Tendongfor, Alan Hubbard, Elvis A. Tanue, Rasheedat Oke, Nahyeni Bassah, Sandra I. McCoy, Larissa Kumenyuy Yunika, Claudia Ngeha Ngu, S. Ariane Christie^3^, Dickson S. Nsagha, Alain Chichom-Mefire, Catherine Juillard.

## References

1. Chang F-R, Huang H-L, Schwebel DC, Chan AHS, Hu G-Q. Global road traffic injury statistics: Challenges, mechanisms and solutions. Chinese Journal of Traumatology. 2020;23(4):216–8.

2. World Health Organization. Global status report on road safety 2018. World Health Organization; 2019.

3. Kwaghgba G, Aondofa T. The effect of economic policies on urban transport patterns: A review of motorcycle usage for urban mobility in Nigeria. Journal of Sustainable Development of Transport and Logistics. 2024;9(2):32–42.

4. Neki K, Mitra S, Wambulwa WM, Job RFS. Profile of low and middle-income countries with increases versus decreases in road crash fatality population rates and necessity of motorcycle safety. Journal of safety research. 2023;84:129–37.

5. Kemajou A, Jaligot R, Bosch M, Chenal J. Assessing motorcycle taxi activity in Cameroon using GPS devices. Journal of transport geography. 2019;79:102472.

6. Wankie C. Epidemiology of crashes and injuries among commercial motorcyclists in Bamenda, Cameroon: A cross-sectional study. University of California, San Diego; 2019.

7. De Rome L, Ivers R, Fitzharris M, Du W, Haworth N, Heritier S, Richardson D. Motorcycle protective clothing: protection from injury or just the weather? Accident Analysis & Prevention. 2011;43(6):1893–900.

8. Wu D, Hours M, Ndiaye A, Coquillat A, Martin J-L. Effectiveness of protective clothing for motorized 2-wheeler riders. Traffic injury prevention. 2019;20(2):196–203.

9. Natarajan G, Rajan T P. Review on the performance characteristics and quality standards of motorcycle protective clothing. Journal of Industrial Textiles. 2022;51(5_suppl):7409S–7427S.

10. Obst M, Rzepczyk S, Głowiński S, Żaba C. Motorbike protective helmets, construction, testing and its influence on the type and severity of injuries of motorbike accident casualties: a literature review. Vibrations in Physical Systems. 2023;34(1).

11. Umar Z, Aslam R, Zaka S. A Review of Global and Local Perspectives on Drivers’ Knowledge, Attitudes, and Practices Towards Road Safety. Bulletin of Business and Economics (BBE). 2024;13(3):429–35.

12. Heydari S, Hickford A, McIlroy R, Turner J, Bachani AM. Road safety in low-income countries: state of knowledge and future directions. Sustainability. 2019;11(22):6249.

13. Nwati MT. The Anglophone Crisis: The birth of warlords, the impact of warlords activity on the people of North West and South West Region of Cameroon. Advances in Applied Sociology. 2020;10(5):157–85.

14. Bang HN, Balgah RA. The ramification of Cameroon’s Anglophone crisis: conceptual analysis of a looming “Complex Disaster Emergency.” Journal of International Humanitarian Action. 2022;7(1):6.

15. Mount Kenya University, A. Senaji FM, Muhonja DrF, Mount Kenya University, Maathai DrR, Mount Kenya University. Association between Utilization of Personal Protective Equipment and Prevalence of Road Traffic Injuries amongst Motorcycle Users in Kibera Constituency, Nairobi County Kenya. International Journal of Current Science Research and Review. 2023;06(02).

16. Alzahrani MM, Alghamdi AA, Alghamdi SA, Alotaibi RK. Knowledge and attitude of dentists towards obstructive sleep apnea. international dental journal. 2022;72(3):315– 21.

17. Nthoki B, Biwott C, Kamau A. Influence of information, education and communication on road safety amongst Boda-boda motorcyclists in Kenyan cities. Eastern African Journal of Humanities and Social Sciences. 2024;3(1):120–30.

18. Corgozinho MM, Montagner MÂ. Sociodemographic profile of motorcyclists and their vulnerabilities in traffic. Revista Brasileira de Medicina do Trabalho. 20(2):262–71.

19. Wankie C, Al-Delaimy W, Stockman J, Alcaraz J, Shaffer R, Hill L. Prevalence of crashes and associated factors among commercial motorcycle riders in Bamenda, Cameroon. Journal of Transport & Health. 2021;20:100993.

20. Rahman MS. VULNERABILITY ASSESSMENT OF MOTORCYCLE RIDING IN DHAKA CITY BY STRUCTURAL EQUATION MODELLING. 2022;

21. Lucci C, Piantini S, Savino G, Pierini M. Motorcycle helmet selection and usage for improved safety: a systematic review on the protective effects of helmet type and fastening. Traffic injury prevention. 2021;22(4):301–6.

22. Ngekeng S, Kibu O, Oke R, Bassah N, Touko DA, Yost MT, Dissak-Delon F, Tendongfor N, Nguefack-Tsague G, Hubbard A, McCoy SI, Christie SA, Chichom-Mefire A, Juillard C. Prehospital factors associated with mortality among road traffic injury patients: analysis of Cameroon trauma registry data. BMC Emergency Medicine. 2024;24(1):194.

23. Liasidis P, Benjamin E, Jakob D, Lewis M, Demetriades D. Injury patterns and outcomes in motorcycle passengers. European Journal of Trauma and Emergency Surgery. 2023;49(6):2447–57.

24. Akbari M, Lankarani KB, Tabrizi R, Vali M, Heydari ST, Motevalian SA, Sullman MJM. The effect of motorcycle safety campaign on helmet use: A systematic review and meta-analysis. IATSS Research. 2021;45(4):513–20.

25. (PDF) Knowledge, Attitude and Compliance with Safety Protective Measures and Devices among Commercial Motorcyclists in Sokoto Metropolis, Northwestern Nigeria. ResearchGate. 2024;

26. Mumuni A, Abeeb Adebayo L, Mynepalli Kameswara Chandra S. Road Use Regulations: Knowledge and Compliance Among Commercial Motorcycle Riders in Oluyole Local Government Area, Ibadan, Nigeria. Journal of Health and Environmental Research. 2023;

27. Mahdavi Sharif P, Najafi Pazooki S, Ghodsi Z, Nouri A, Ghoroghchi HA, Tabrizi R, Shafieian M, Heydari ST, Atlasi R, Sharif-Alhoseini M. Effective factors of improved helmet use in motorcyclists: a systematic review. BMC public health. 2023;23(1):26.

28. Fondzenyuy SK, Fowo Fotso CS, Feudjio SLT, Usami DS, Persia L. Self-Reported Speed Compliance and Drivers Speeding Behaviour in Cameroon. Future Transportation. 2024;4(2):659–80.

29. Akande AT, Rodríguez Parrón M, Jariot M, Martínez M. Risky Driving Attitudes and Behaviours among Commercial Drivers and the Rate of Accidents on Nigerian Roads: A Case Study of Abuja and Lagos State. 2021.

30. Urréchaga EM, Kodadek LM, Bugaev N, Bauman ZM, Shah KH, Abdel Aziz H, Beckman MA, Reynolds JM, Soe-Lin H, Crandall ML, Rattan R. Full-face motorcycle helmets to reduce injury and death: A systematic review, meta-analysis, and practice management guideline from the Eastern Association for the Surgery of Trauma. The American Journal of Surgery. 2022;224(5):1238–46.

31. Dewa A. The Influence of Rider Concentration, Vehicle Completeness and Traffic Discipline on Motorcycle Rider Safety. Research Horizon. 2023;3(4):391–400.

32. Dada OT, Fasina SO, Agbabiaka HI, Salisu UO, Ogunseye NO, Olawale OA. Occupational hazards and risks among commercial motorcyclists in the peri-urban city of Lagos, Nigeria. International Journal of Occupational Safety and Ergonomics. 2022;

